# Single Cell Sequencing of Human Langerhans Cells Identifies Altered Gene Expression Profiles in Patients with Atopic Dermatitis

**DOI:** 10.1101/2024.05.06.24306801

**Authors:** Sara M. Tamminga, M. Marlot van der Wal, Elise S. Saager, Lian F. van der Gang, Celeste M. Boesjes, Astrid Hendriks, Yvonne Pannekoek, Marjolein S. de Bruin, Femke van Wijk, Nina M. van Sorge

## Abstract

Atopic dermatitis (AD) is characterized by dysregulated T cell immunity and skin microbiome dysbiosis with predominance of *Staphylococcus aureus* (*S. aureus*). Emerging evidence suggests a role for *S. aureus* in exacerbating AD skin inflammation. We have previously shown that specific glycosylation of *S. aureus* cell wall structures amplifies skin inflammation through interaction with Langerhans cells (LCs). However, the role of LCs in AD remains poorly characterized. Here, we performed single cell RNA-sequencing of primary epidermal LCs and dermal T cells isolated from skin biopsies of AD patients and healthy controls, alongside specific glycoanalysis of *S. aureus* strains isolated from the AD lesions. Our findings reveal four LC subpopulations, including two steady-state clusters (LC1 and LC1_H_) and two pro-inflammatory/matured subsets (LC2 and migratory LCs). The latter two subsets were enriched in AD skin. AD LCs showed enhanced expression of C-type lectin receptors, the high-affinity IgE receptor (FcεR1), and activation of prostaglandin and leukotrienes biosynthesis pathways, as well as upregulated transcriptional signatures related to T cell activation pathways and increased expression of CCL17 (specifically LC2) compared to healthy LCs. Correspondingly, T helper 2 and regulatory T cell populations were increased in AD lesions. Our study provides proof-of-concept for a role of LCs in connecting the *S. aureus*-T cell axis in the AD inflammatory cycle.

## Introduction

Atopic dermatitis (AD) or eczema is a chronic inflammatory skin disorder that affects at least 230 million people worldwide (Hay et al., 2014). Patients with AD suffer from chronic inflamed skin lesions, which can severely decrease quality of life. AD is a multifactorial disease, characterized by dysregulated immunity, compromised skin barrier function, and a dysbiotic skin microbiome composition with predominant presence of the bacterium *Staphylococcus aureus* (*S. aureus*) (Callewaert et al., 2020, Eyerich and Novak, 2013, Fyhrquist et al., 2019). These components also impact each other to enhance and perpetuate the chronic inflammatory nature and barrier defects that characterize AD skin, making it challenging to dissect cause from consequence.

In AD, IL4 and IL13 are key drivers of the skewed T helper 2 (Th2)-mediated inflammation. Consequently, dupilumab and tralokinumab, two human monoclonal antibodies that interfere with IL4/IL13 responses, rapidly improve skin physiology and microbial ecology in AD patients (Beck et al., 2014, Sander et al., 2024). Importantly, AD disease severity, especially flares, correlate with decreased microbial diversity and dominant *S. aureus* colonization in lesions (Callewaert et al., 2020, Khadka et al., 2021, Saheb Kashaf and Kong, 2024). Based on human epidemiological, human microbiome, and experimental mouse studies, a view has emerged of a *S. aureus*-induced inflammatory amplification loop in AD skin; individuals with a skin barrier defect become colonized by *S. aureus* strains that have particular pro-inflammatory traits, and reciprocally these strains are better equipped to colonize inflamed skin (Byrd et al., 2017, Geoghegan et al., 2018, Iwamoto et al., 2019). However, how lesional *S. aureus* colonization affects the local immune response and the overall disease pathogenesis remains unclear.

Glycosylation of *S. aureus* cell wall components affects interaction with specific immune components including human Langerhans cells (LCs) (Hendriks et al., 2021b, van Dalen et al., 2019). LCs are key sentinel cells in the skin epidermis, which are characterized by the expression of CD1a and langerin (CD207). Human langerin specifically recognizes β-linked-N-acetylglucosamine (β-GlcNAc) moieties linked to surface-expressed wall teichoic acid (WTA) molecules of *S. aureus*. Presence of WTA-β-GlcNAc depends on staphylococcal *tarS* and *tarP* genes and results in enhanced LC maturation and production of pro-inflammatory cytokines (Hendriks et al., 2021b, van Dalen et al., 2019). In contrast, modification of *S. aureus* WTA with α-GlcNAc (dependent on *tarM*) reduces LC interaction and subsequent cytokine production (Hendriks et al., 2021b, van Dalen et al., 2019). Overall, these data linked specific *S. aureus* characteristics to inflammatory processes in the skin. Importantly, LCs can directly activate skin-resident T memory cells or migrate from the epidermis to the lymph nodes to activate naïve T cells (Romani et al., 2012). Indeed, *S. aureus-*specific T cells, which predominantly secreted Th2-related IL4 and IL13, were found in AD patients (Farag et al., 2022, Hendriks et al., 2021a). Additionally, it was shown that human T cells can be activated by a CD1a-presented *S. aureus* membrane phospholipid, resulting in production of Th2 cytokines (Monnot et al., 2022). The observation that these T cells are expanded in AD patients (Monnot et al., 2022), supports the concept that LCs may connect *S. aureus* detection in the epidermis and T cell activation in the dermis, thereby acting as critical players in the initiation and perpetuation of skin inflammation. This could particularly occur at AD lesional sites that are heavily colonized with *S. aureus*.

In this study, we performed single cell RNA-sequencing of primary epidermal LCs and dermal T cells, isolated from skin biopsies of AD patients and healthy controls, and analyzed their transcriptional profiles. Complementary, we obtained colonizing *S. aureus* strains through skin swabs of the same AD lesions and determined the specific WTA glycoprofile to analyze potential relation to the LC transcriptional response. Overall, our results provide proof-of-concept that LCs may perpetuate the AD inflammatory cycle by bridging *S. aureus* recognition and T cell activation.

## Results

### Single cell RNA sequencing reveals four human Langerhans cell clusters

We sorted viable CD45+ HLA-DR+ CD1a+ CD207+ human LCs from the epidermis of healthy skin (n=3) and AD lesional skin (n=4; gating strategy in Figure S1). The LC yield for each donor is summarized in Table S1. Single cells were submitted for RNA sequencing and after quality control, dimensionality reduction, and clustering, the pooled 1,677 high-quality LCs separated into four LC clusters (Figure 1A). Each cluster was distinguished based on differentially-expressed genes (DEGs) (Figure 1B; Table S2). LC subset 1 (LC1, 830/1,677 cells) was characterized by high expression of classical LC markers langerin (CD207, Figure 1B, C) and CD1a (Figure 1D), the CCR4 ligand CCL22, and the actin cross-linking protein MARCKS, which is involved in regulation of cell migration (Figure 1B). Using gene set enrichment analysis (GSEA), antigen processing and presentation pathways emerged as the most enriched pathways in this cluster (Figure 1I). In contrast, non-classical antigen-presenting receptors CD1b and CD1c showed low expression (Figure 1E, F). In addition to LC1, the other large LC subset was designated LC1_H_ (Figure 1A, 711/1,677 cells). This cluster showed overlapping characteristics with LC1 (Table S3, Figure 1C-H), except for the high expression of heat shock proteins (HSPs), including HSPA6, DNAJB1 and HSHPH1 (Figure 1B, Table S2,3). HSPs prevent aggregation of denatured proteins and cluster to pathways associated with stress (heat, cellular stress, and unfolded proteins) (Figure 1J). The remaining LC subsets, LC2 and migratory LCs, contained relatively few cells (62 and 74 cells, respectively) compared to LC1 and LC1_H_. LC2 showed high expression of molecules involved in host defense: lysozyme (LYZ) and C1QB, the first component of the serum complement system (Figure 1B). Furthermore, molecules involved in antigen uptake and presentation including CLEC10A (MGL), CD1b, and CD1c, were upregulated in this cluster (Figure 1B, E, and F). Correspondingly, immune defense response pathways were enriched in the LC2 subset (Figure 1K). Finally, the migratory LC cluster included an enrichment of upregulated genes involved in cell motility and migration such as: FSCN1, MARCKSL1 and LSP1 (Figure 1B, Table S2). Other upregulated genes included BIRC3, LAMP3, IL7R and IDO1 (Figure 1B, Table S2). LC maturation and activation marker CD83 (Figure 1G) as well as CCR7 (Figure 1H), which is needed for migration towards lymph nodes, were both highly expressed in this cluster whereas langerin (CD207) and CD1a were downregulated (Figure 1C, D). The pattern of up- and downregulated genes did not aggregate into specific enriched pathways for this cluster. Based on the gene expression in these four clusters, the LC1 and LC1_H_ subpopulations appear to be more ‘steady-state’ LCs, whereas LC2 and migratory LCs appear to be more pro-inflammatory and matured LCs, respectively.

**Figure 1.**
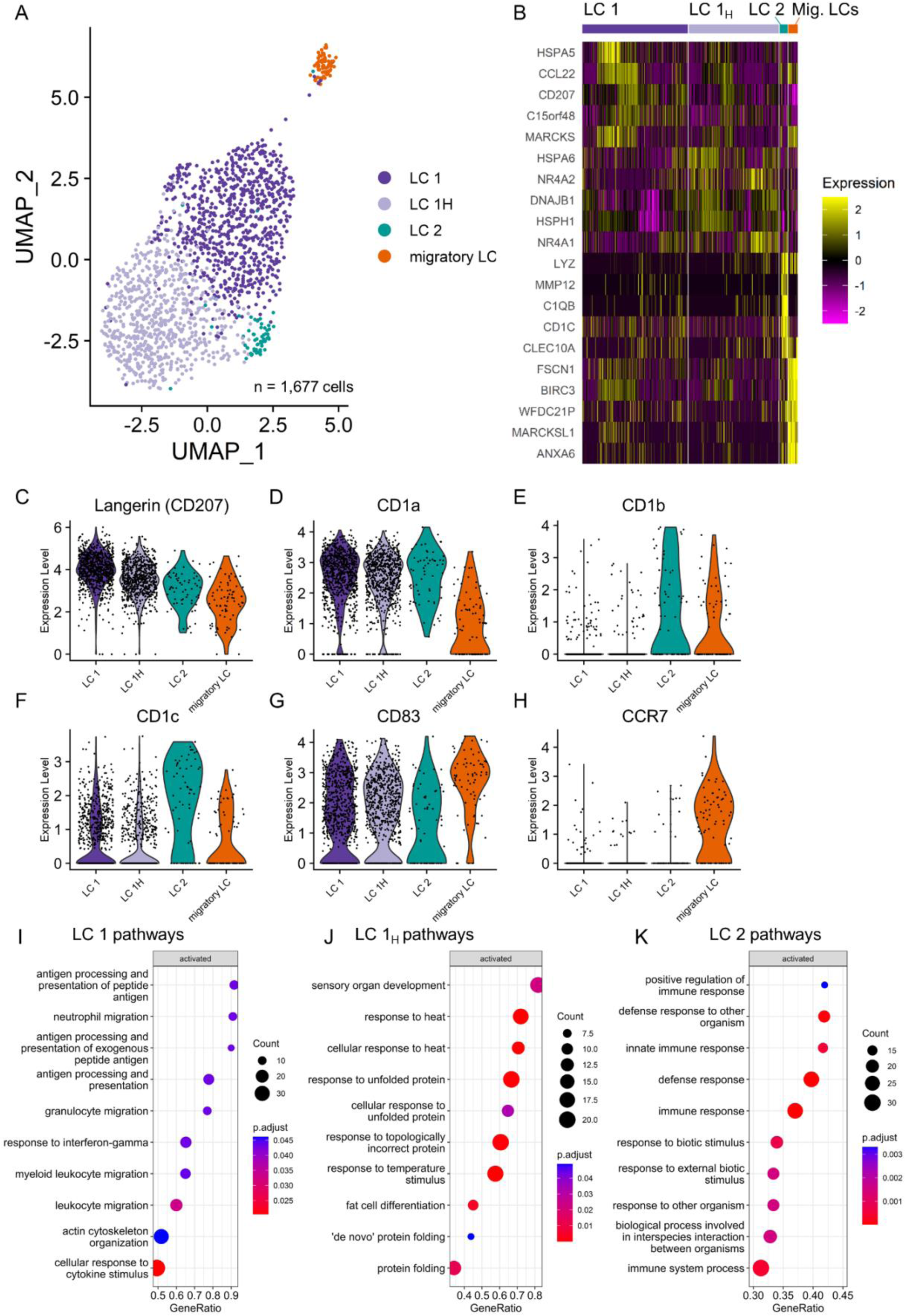
Identification and transcriptional profile of four primary Langerhans cell clusters in human skin tissue. (A) Single cell RNA sequencing Uniform Manifold Approximation and Projection (UMAP) plot of 1,677 individual LCs from the epidermis of healthy and AD individuals, colored by cluster. (B) Heatmap of top 10 upregulated differentially expressed genes (DEG) per cluster. (C-H) Violin plots showing expression of indicated LC surface markers per LC subcluster. (I-K) Pathway analysis for subpopulations (I) LC 1, (J) LC1_H_ and (K) LC 2. Only 10 pathways with highest normalized enrichment score (NES) are shown. Diameter of circle indicates the total number of core enrichment genes. GeneRatio is the number of core genes divided by the total number of genes in the pathway. Color indicates the adjusted *P* value corrected for gene set size and multiple hypothesis testing.

### Different LC cluster abundance and LC gene expression in epidermal lesional AD skin versus healthy skin

We assessed the relative abundance of the identified LC clusters in AD patients and healthy controls (Figure 2A). Across all individuals, LC1 and LC1_H_ were the most abundant, collectively constituting over 85% of isolated LCs (Figure 2A, Figure S2). Interestingly, proportions of minor clusters LC2 and the migratory LCs were enriched in samples from AD lesional skin compared to skin from healthy controls (Figure 2A, permutation test, FDR = 0.0056 and 0.00040, respectively). Next, we analyzed whether LCs from AD lesions had different gene expression signature compared to LCs from healthy skin. DEG analysis comparing all LCs from AD patients to all healthy control LCs identified 160 upregulated genes and 174 down regulated genes (Figure 2B, Table S4). TNF-α related genes CD40, TRAF1, and NFKBI were downregulated in AD LCs (Figure 2B). Upregulated genes in LCs from AD patients included TGF-β response genes: TACSTD2 and TGFBI, which are involved in the inhibition of cell adhesion (Figure 2B). Correspondingly, migration and motility-related genes were upregulated in AD LCs including GSN, ACTB, CAP1, ARPC1B and VIM (Figure 2B). Other genes of interest that were upregulated in AD LCs included ALOX5, which catalyzes the first two steps in the biosynthesis of leukotrienes, and the high-affinity IgE receptor (FcεR1A, Figure 2B). Based on our findings in the DEG analysis, we investigated three gene signatures based on literature: FcεR1 signaling, prostaglandin and leukotriene biosynthesis, and C-type lectin receptor signaling (Table S5). We scored the single cells for all three gene-sets, based on their gene expression profiles. The expression of genes involved in FcεR1 receptor signaling (Figure 2C), biosynthesis and signaling of prostaglandins and leukotrienes (Figure 2D), and signaling of C-type lectins (Figure 2E), was increased in all LC subsets from AD patients compared to healthy controls. Subsequently, we analyzed the pattern of up- and downregulated genes in AD LCs compared to healthy LCs (Table S4) with GSEA using a comprehensive collection of pathways sourced from the Pathway Interaction Database (Schaefer et al., 2009). Correspondingly, GSEA revealed activation of the FcεR1 pathway in LCs from AD skin lesions and suppressed TNF- and CD40/CD40L signaling pathways (Figure 2F). Furthermore, the RAC1 pathway was activated (Figure 2F), which orchestrates actin polymerization and lamellipodia formation, for actin-based motility and reorganization of the cytoskeleton (Schaefer et al., 2009). Finally, several pathways associated with T cell activation, i.e. the Calcium Signaling in the CD4+ TCR pathway, the CD8+ TCR Downstream pathway and the Calcineurin-dependent NFAT Signaling In Lymphocytes pathway, were found to be activated (Figure 2F). Subsequently, we specifically analyzed DEGs in the migratory LCs and LC2, which were enriched in the AD lesions (Figure 2A). In migratory LCs from AD patients, NET1, which is involved in cell motility, was upregulated (Figure 3A). Intriguingly, in AD LC2, the C-type lectin receptor CLEC10A (MGL) was upregulated, as was the AD biomarker and Th2 cell attractant CCL17 (TARC) (Figure 3B). These findings indicate that there are differences in the proportion of LC subsets as well as gene expression profiles of LCs isolated from AD lesions compared to LCs from healthy individuals.

**Figure 2.**
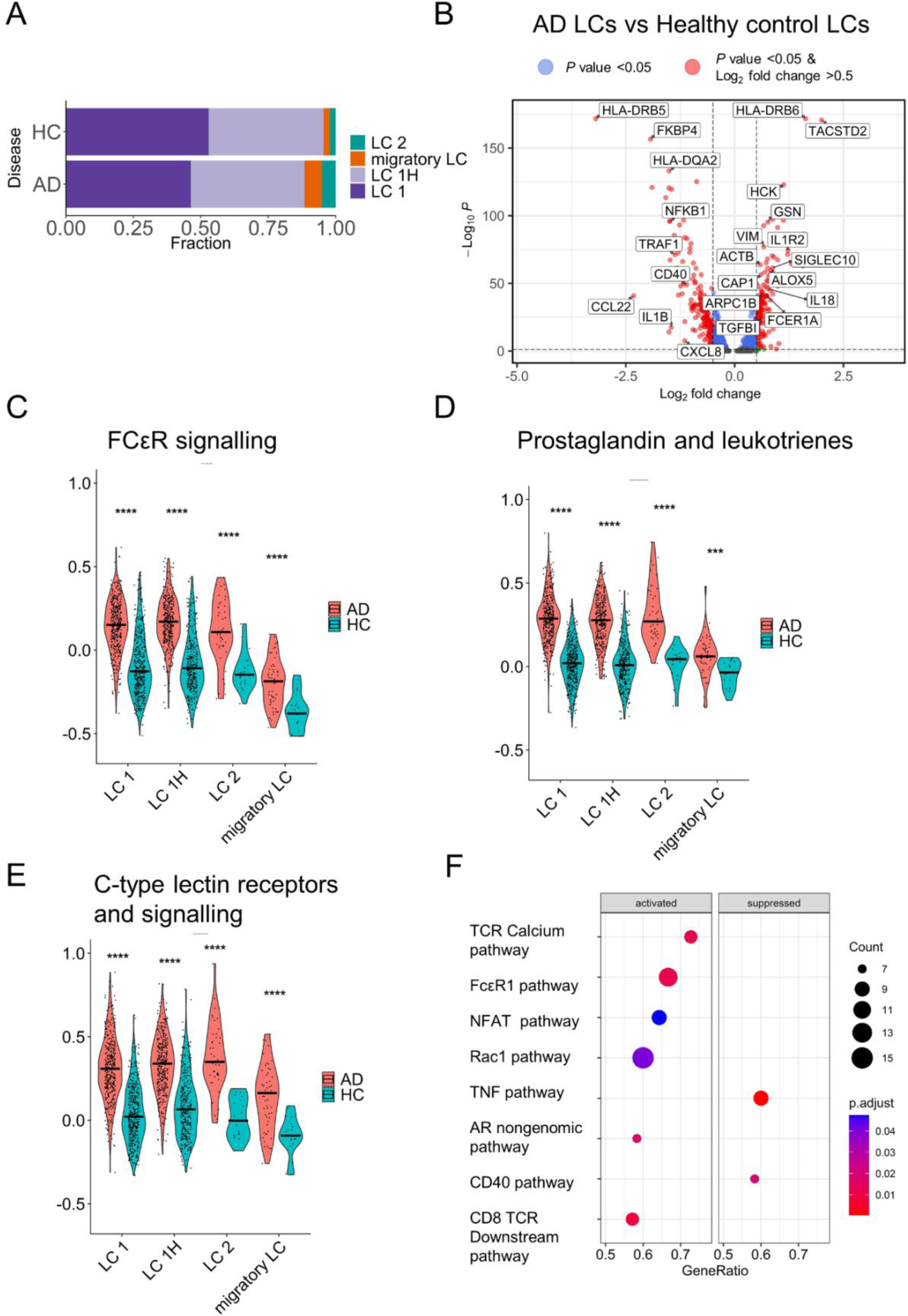
Divergent abundance and differentially-expressed genes of LCs from AD patients versus healthy controls. (A) Mean distribution of the four LC clusters in the epidermis of healthy individuals (HC; n=3) and AD patients (AD; n=4). Proportion of LC 2 and migratory cells was higher in AD than in HC skin (permutation test, *P* = 0.0009 and *P* = 0.0001, False Discovery Rate (FDR) = 0.0056 and 0.00040, respectively. (B) Volcano plot indicating DEGs in all AD LCs compared to all LCs from healthy controls. Genes that reach Log_2_ Fold Change cut-off >0.5 and adjusted *P* value cut-off <0.05 are depicted as red circle. (C-E) Violinplots of the module scores for (C) Fc ε Receptor signaling-, (D) prostaglandin and leukotrienes- and (E) C-type lectin receptors and signaling genesets. Wilcoxon rank sum test was performed on module scores comparing AD vs healthy controls (HC) for each LC cluster; **** *P* < 0.0001, *** *P* < 0.001. (F) Pathway analysis of AD LCs, top 8 activated or suppressed pathways with lowest *P* values are displayed. Diameter of circle indicates the total number of core enrichment genes. GeneRatio is the number of core genes divided by the total number of genes in the pathway. Color indicates the adjusted *P* value corrected for gene set size and multiple hypothesis testing.

**Figure 3.**
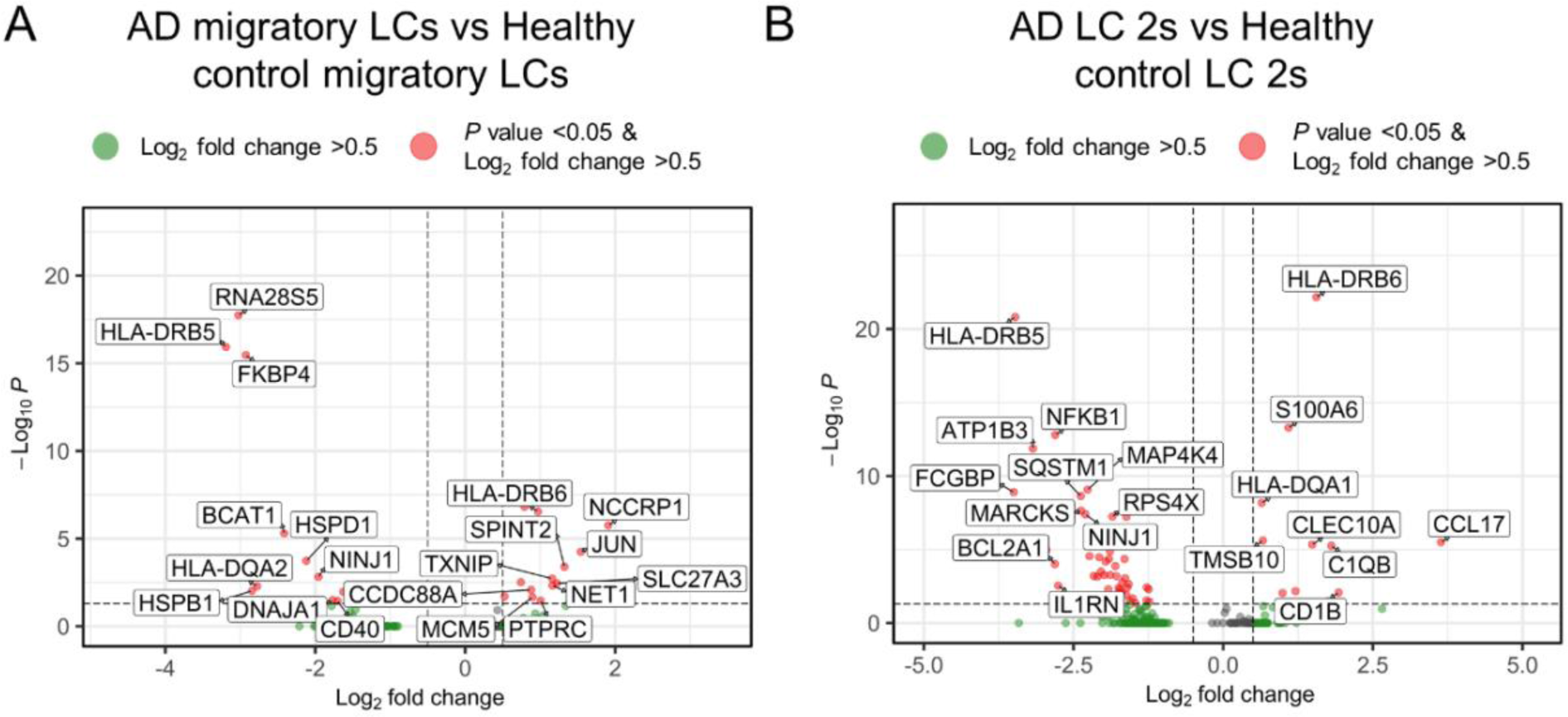
Differentially-expressed genes of migratory LCs and LC2 subcluster from AD patients versus healthy controls. Volcano plot indicating DEGs in AD subcluster LC 2 (A) and AD subcluster migratory LCs (B) compared to healthy control LCs from corresponding subcluster. Genes that reach cut-off values of Log_2_ Fold Change >0.5 and adjusted *P* value <0.05 are depicted as red circle.

### Dermal T cell subset abundance and differential gene expression in lesional AD versus healthy control skin

The enrichment of T cell activation pathways in AD LCs, as well as the increased expression of CCL17 (TARC) in LC2 of AD patients, prompted us to investigate the transcriptional profile of CD45+ CD3+ sorted T cells isolated from the dermis of the same donors (except for AD04, due to technical limitations). Figure S3 shows the gating strategy for the T cell sort. T cell yield for each donor is summarized in Table S1, amounting to a total of 1,719 single dermal T cells after quality control. Single T cells were clustered with a resolution of 0.7 (Figure S4A). Based on low total RNA molecules (nCount_RNA), low unique RNA molecules (nFeature_RNA), and the lack of significant or meaningful DEGs, cluster 3 and 7 were excluded from further analysis (Figure S4B, C). A total of 1,475 dermal T cells divided over six distinct clusters remained (Figure 4A), clusters were re-named based on the expression of general T cell markers and DEGs (Figure 4B, Figure S4C, Table S6). Cluster “CD8” co-expressed CD8A and CD8B (Figure S4). DEGs in the CD8 cluster included the effector chemokine CCL5, granulysin (GNLY), and granzyme K (GZMK) (Figure 4B, Table S6). We also identified a cluster (“HSP”) with low CD4 and CD8 expression, but upregulation of HSPs (Figure 4B, Table S6). Furthermore, four CD4-expressing clusters were identified: ‘CD4_A_’, ‘CD4_B_’, ‘CD4 Th2’ and ‘Tregs’. Cluster CD4_A_ expressed lymphotoxin β (LTB) among its DEGs (Figure 4B, Table S6). Cluster CD4_B_ showed DEGs related to cell-motility and chemotaxis including ACTB, ACTG1 and S100A4 (Figure 4B, Table S6). Cluster CD4 Th2 was named based on expression of CXCR4, IL7R and GATA3 (Figure 4B, Table S6). Finally, a Treg cluster was identified characterized by the top DEGs: FOXP3, CTLA4 and IL2RA (Figure 4B, Figure S4, Table S6). The larger clusters CD4_A_, CD4_B_ and HSP, collectively constituted over 67% of all T cells (n=1,000) and were evenly distributed across individuals, independent of disease status (Figure 4C, Figure S5). In contrast, the smaller clusters, consisting of CD8 T cells (n=186), CD4 Th2 (180) and Tregs (n=109), showed more variability (Figure 4C). CD8+ T cells were predominant in healthy controls compared to AD patients (permutation test, FDR = 0.0033), whereas CD4 Th2 and Tregs were more prevalent in AD patients (Figure 4C, permutation test FDR= 0.032 and 0.0033, respectively).

**Figure 4.**
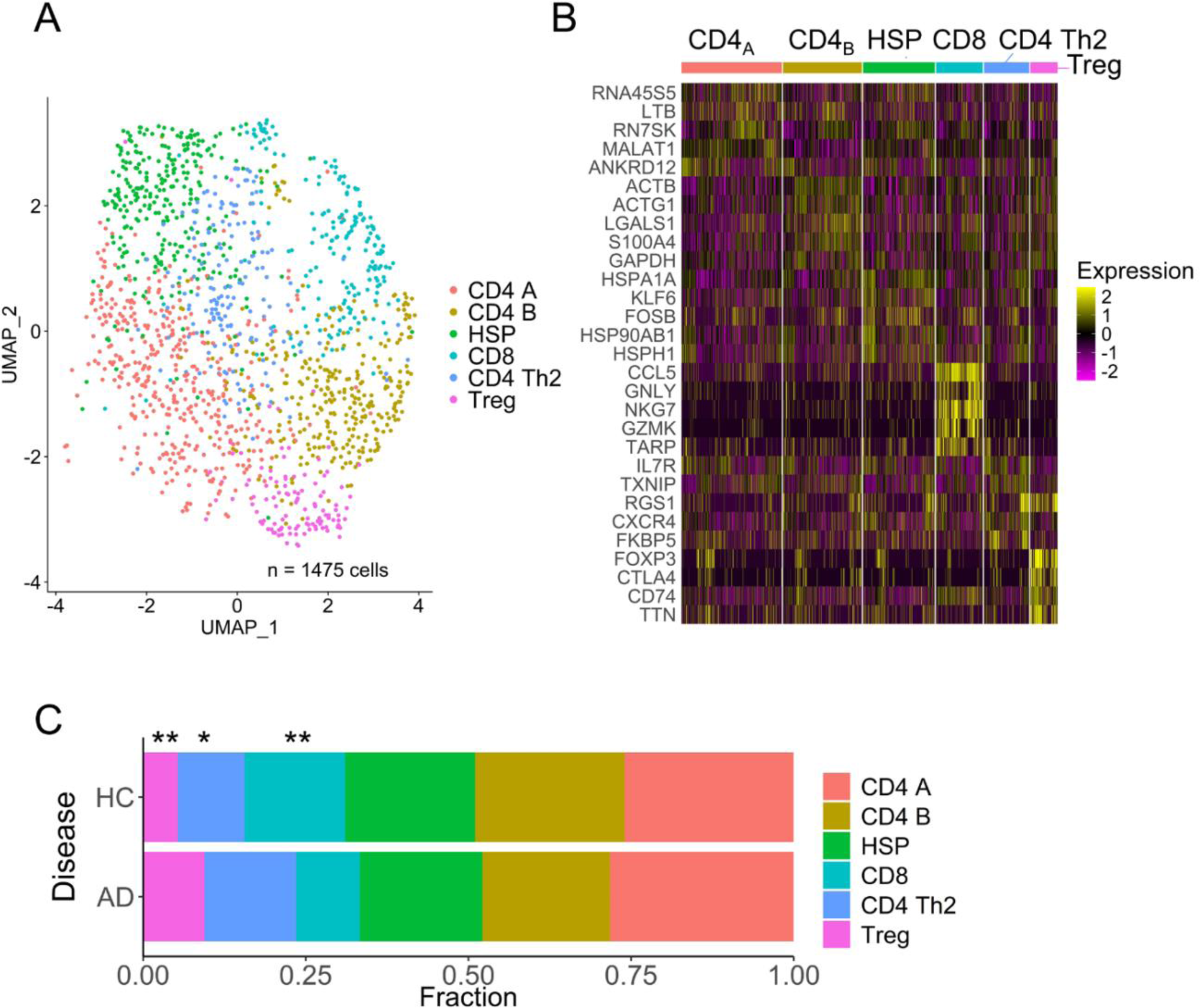
Differences in abundance of T-cell clusters in AD patients versus HCs. (A) UMAP plot of 1,475 individual CD3+ T cells from healthy and AD, colored by cluster. (B) Heatmap of top 5 upregulated differentially expressed genes (DEG) per cluster. (C) Distribution of six T cell clusters in three healthy individuals (HC) and three AD patients. Proportion of CD8 T cells was lower, and CD4 Th2 cells and Tregs was higher in AD than in HC skin (permutation test, *P* = 0.0006, *P =* 0.016 and *P* = 0.0011, False Discovery Rate (FDR) = 0.0033, 0.032 and 0.0033, respectively).

### AD colonizing *S. aureus* strains only express β-GlcNAc WTA

*S. aureus* was recovered from three out of four individuals (Table 2). Of note, the patient without *S. aureus* culture (AD02) exhibited the lowest EASI score (EASI: 6.2, Table 1) and the lowest proportion of LC2 and migratory LCs (Figure 2A). Since we previously demonstrated that *S. aureus* WTA glycosylation can impact LC inflammatory responses (Hendriks et al., 2021b, van Dalen et al., 2019), we assessed the presence of the different WTA glycosyltransferase genes *tarS, tarM*, and *tarP* in the recovered isolates. Interestingly, our analysis revealed the presence of *tarS* and *tarP* and absence of *tarM* in all three AD isolates (Table 2). We confirmed the presence of β-GlcNAc and absence of α1,4-GlcNAc using specific Fab fragments for flow cytometric analysis (Table 2). These observations are of interest since β-GlcNAc WTA is related to an inflammatory LC phenotype, whereas α-GlcNAc WTA decorations dampen this response. These findings therefore underline the possibility that the differential gene expression profiles of AD LCs, may contribute to the observed inflammatory response in AD skin lesions.

**Table 1.** Patient characteristics at moment of sampling.

| Clinical Characteristics | Measure |
| --- | --- |
| Age, y, median (IQR) | 34 (24-45) |
| Male, n (%) | 3 (75%) |
| Atopic comorbidities (n, %) |  |
| Allergic asthma | 2 (50) |
| Allergic rhinitis | 3 (75) |
| Allergic conjunctivitis | 1 (25) |
| Food allergy | 1 (25) |
| EASI score, median (IQR) | 8.6 (6.2-11) |
| IGA score, median (IQR) | 2.0 (2.0-2.3) |
| Weekly average pruritus NRS, median (IQR) | 7.0 (4.0-10) |

Abbreviations: AD, atopic dermatitis; EASI, Eczema Area Severity
Index; IQR, interquartile range; NRS, numeric rating scale.

**Table 2.** Overview of *S. aureus* colonization status, *tar* gene presence and glycoprofile.

| **Patient** | ***S. aureus* positive culture** | ***tar* gene presence** | **WTA glycoprofile** |
| --- | --- | --- | --- |
| AD01 | Yes | *tarS, tarP* | β-GlcNAc |
| AD02 | No | - | - |
| AD03 | Yes | *tarS, tarP* | β-GlcNAc |
| AD04 | Yes | *tarS* | β-GlcNAc |

## Discussion

AD pathogenesis is characterized by immune dysregulation, specifically a type 2 T cell response, and an altered skin microbiome marked by highly enriched *S. aureus* colonization. Despite the emerging perception for a *S. aureus*-associated inflammatory amplification loop in AD, our understanding on the molecular pathways and the host immune interactions underlying this phenomenon remains limited. Using single cell RNA-sequencing of primary epidermal LCs, and dermal T cells isolated from skin biopsies of AD patients and healthy controls, we demonstrated that similar to T cell-, also LC subset composition and transcriptome signatures differed between AD patients and healthy controls. In addition, we identified the presence of *S. aureus* in 3 out of 4 AD patients, all of which expressed glycosylated WTA that enhances LC inflammatory responses. Overall, our results suggest a central role for LCs in connecting the *S. aureus*-T cell axis in AD.

Using single cell RNA sequencing, we identified four LC clusters in the skin epidermis. Three of our subsets correspond to LC subsets recently described in human foreskin (Liu et al., 2021). Specifically, the transcription profiles of our migratory LC and LC1 clusters largely correspond to the respective population of CCR7^+^ migratory LCs and a Langerin^+^, CD1a-expressing steady-state cluster (named LC1) of Liu *et al*. In addition, they reported the presence of a LC2 subpopulation, which had CLEC10A (MGL) and lysozyme (LYZ) among its top DEGs and appears similar to our LC2 cluster, although their LC2 cluster was larger in number. Our fourth LC subset, HSP-enriched LC1_H_ cluster, was not observed by Liu *et al*. Conversely, they report a small active LC cluster, which we did not identify in our data. These discrepancies, as well as the smaller proportion of LC2 may be explained by the difference in sampled body site, i.e. trunk and extremities versus foreskin, which is in line with the previous report that LC2 was more prominent in foreskin but absent in trunk skin (Liu et al., 2021).

We observed an enrichment of migratory LCs and LC2 in AD lesional skin compared to healthy skin. Interestingly, Liu *et al*. reported increased proportions of corresponding LC subsets in psoriasis lesional skin (Liu et al., 2021). These findings may suggest that the increased abundance of these subsets is relevant in the pathogenesis of inflammatory skin diseases. As psoriasis and AD exhibit distinct immunological characteristics and dysregulated pathways, the specific role of these LC clusters in both diseases remains to be determined.

Our study revealed increased numbers of Tregs in AD patients’ skin versus healthy controls, corresponding to previous reports (Hanafusa et al., 2013, Roesner et al., 2015, Yasunori Ito, 2009). Moreover, previous research suggested a potential role for LCs in promoting Treg expansion *in vivo* (Seneschal et al., 2012, Szegedi et al., 2009). Possibly, this expansion is associated to increased expression of CLEC10A in the LC2 cluster of AD patients; since CLEC10A induced IL10 expression induces Tregs (Geijtenbeek and Gringhuis, 2016, van Vliet et al., 2013). Whether increased Treg populations are a response to increased inflammation or contribute to AD pathogenesis, is currently unresolved.

*S. aureus* was cultured from skin swabs of 3 out of 4 AD patients. Combining flow cytometry and PCR analysis, we determined that all cultured *S. aureus* isolates expressed β-GlcNAc-WTA, while α-GlcNAc-WTA was absent. It is known that certain *S. aureus* clonal complexes are overrepresented in AD lesions (Fleury et al., 2017, Rojo et al., 2014, Shoko Obata, 2023, Yeung et al., 2011), and that specific *S. aureus* glycoforms are associated with certain clonal complexes (Tamminga et al., 2022). Intriguing in this respect is the observation that CC30 strains, which express α-GlcNAc producing TarM, are generally underrepresented, while CC1 strains are over-represented in AD versus healthy skin (Fleury et al., 2017, Rojo et al., 2014, Shoko Obata, 2023, Yeung et al., 2011). Whether the presence or absence of β- or α-GlcNAC-WTA is related to colonization of AD lesions, needs to be studied in larger cohorts of *S. aureus* strains that are isolated from AD skin.

*S. aureus* exposure may also contribute to the induction of other inflammatory markers, such as AD-biomarker CCL17 (TARC) (Katsuhiko Matsui, 2014). This chemokine was highly upregulated in LCs from AD patients, specifically in the expanded LC2 subset. Interestingly, dupilumab treatment in AD patients resulted in reduction of serum CCL17, which correlated with significant reduction in *S.* aureus abundance on lesional skin (Simpson et al., 2023). Similarly, we observed upregulation of ALOX5 and the corresponding leukotriene biosynthesis pathway in AD LCs, which can be induced by *S. aureus* exotoxins in neutrophils (Romp et al., 2020). Both findings may suggest that in AD lesional skin, *S. aureus* interacts with epidermal LCs, thereby inducing CCL17 and leukotriene production and aggravating AD.

A limitation of the study is the small number of samples, i.e. 4 biopsies from AD patients and 3 from healthy controls. In addition, we focused only on LCs and T cells, based on their superficial location and described interaction with *S. aureus* and importance in AD pathogenesis, respectively. This excludes the predominant cell type in the skin, i.e. keratinocytes, which may also contribute to AD pathogenesis and could affect the transcriptome of studied immune cells. Nonetheless it is the first study to reveal a potential role for human LCs as potential links between *S. aureus* exposure and increased disease activity in AD patients. To increase our understanding of the interplay between LCs, *S. aureus* and T cells in the context of healthy and diseased skin, future studies should include other skin cells, increase the number of sequenced cells and include more AD patients and healthy controls. This knowledge could potentially lead to the development of interventions that specifically target LC activity, to break the inflammatory cycle at an early stage, providing new therapeutic strategies for managing inflammatory skin disorders like AD.

## Materials and methods

### Patient inclusion

This study included adult patients (n = 4) with mild to severe AD (Table 1, Table S7), treated at the Department of Dermatology and Allergology in the National Expertise Center for Atopic Dermatitis, University Medical Center Utrecht, The Netherlands. The included patients did not receive systemic treatment over the past 6 months. Punch biopsies (3 mm, 1 per patient) were taken from lesional skin to single cell sort T cells and LCs for RNA-sequencing. All included patients gave their consent according to our biobank protocol, which has been approved by the Medical Ethical Committee of the University Medical Center Utrecht (approval number 12-407). Abdominal or breast *ex vivo* human skin (healthy controls, n=3), was obtained from local hospitals after cosmetic surgery. The procedure was according to the ethical principles of the Declaration Helsinki.

### Sample collection skin swabs

Prior to disinfection and punch biopsy, the skin lesions of the AD patients were swabbed to collect resident microbiota. Sterile flocked swabs were wetted with a buffer (phosphate-buffered saline, PBS; with 0.05% Tween®20, Sigma) and rubbed for 30 seconds on lesional skin. The swabs were stored in sterile tubes and immediately snap frozen in liquid nitrogen, and thereafter stored at −80 °C until further analysis.

### *S. aureus* colonization status on AD lesions and WTA glycoprofile characterization

The collected skin microbiota swab was selectively cultured to determine the presence of *S. aureus.* Each swab was streaked on Tryptic soy agar supplemented with 5% sheep red blood cells (Becton, Dickinson) and grown overnight at 37°C. When possible, up to 4 single gold-colored colonies per patient were re-streaked and grown overnight. *S. aureus* identity was confirmed using MALDI-TOF/MS (Bruker). Gene presence of *tar-*glycosyltransferases (*tarS, tarP* and *tarM*) was checked for cultured *S. aureus* isolates by PCR (primers are listed in Table S8). *S. aureus* WTA glycosylation was confirmed using WTA-specific Fab fragments as previously described (Hendriks et al., 2021b, Tamminga et al., 2022). Briefly, bacteria were grown to mid-exponential phase from overnight cultures in Tryptic Soy Broth (Oxoid), at 37°C with agitation, collected and resuspended at an optical density at 600 nm (OD_600_) of 0.4 in PBS (pH = 7) with 0.1% BSA (Sigma). Next, bacteria were incubated with monoclonal Fab fragments specific to β-GlcNAc (clone 4497; 3.3 μg/mL) or α1,4-GlcNAc WTA (clone 4461; 3.3 μg/mL). After washing in PBS 1% BSA, goat F(ab’)2 anti-human Kappa AF647 (Southern Biotech #2062-31) was used to detect bound Fab Fragments. Fluorescence was determined by flow cytometry on a BD FACSCanto II Flow Cytometer (BD Bioscience) and data were analyzed using FlowJo version 10.

### Enzymatic digestion of skin biopsies

Full-thickness biopsies of lesional and healthy control skin were collected in an Eppendorf tube containing ice-cold 1.5 mL DMEM with glutaMAX (Gibco, 31966-02), 10% fetal bovine serum (FBS, Invitrogen), 100 U/mL penicillin-streptomycin (Gibco, 115288876), and were processed immediately. After washing in sterile PBS, skin biopsies were incubated in RPMI1640 with an enzyme mix for 16 hours at 4⁰C with constant rotation according to the Miltenyi Epidermis Dissociation Kit. The next day, epidermis and dermis were separated, and both were fully digested to single cell suspension using the Miltenyi Epidermis Dissociation Kit or the Miltenyi Whole Skin Dissociation Kit, respectively.

### Cell staining and SORT-seq

To prepare samples for fluorescence-activated single cell sorting, the single cell suspensions of the epidermal and dermal skin cells were washed in PBS. To determine viability, eBioscience Fixable Viability Dye eFluor 506 (Invitrogen, 65-2860-40) was used. Next, each single cell suspension was incubated for 25 minutes at 4°C with antibody mix (Table S9). For each donor, live CD45+ HLA-DR+ CD1a+ Langerin+ LCs from the epidermal skin, and CD45+CD3+ T cells from the dermal skin were sorted into hard shell 384-well plates (Biorad) using a FACSAria™ III (BD; gating strategy in Figure S1 and S4). Plates contained 5 μl of vapor-lock (Qiagen) with 100-200 nl of primers, dNTPs and synthetic mRNA SpikeIns and were immediately centrifuged (2,000 x *g*) and frozen at −80⁰C. Cells were prepared for SORT-seq as previously described (Muraro et al., 2016). Illumina sequencing libraries were prepared with the TruSeq small RNA primers (Illumina) and sequenced single end at 75-bp read length with 75,000 reads per cell on a NextSeq500 platform (Illumina). Sequencing reads were mapped against the reference human genome (GRCh38) using Burrows-Wheeler Alignment tool (BWA).

### Clustering of Single Cell RNA sequencing data

We analyzed the unique molecular identifiers (UMI)-corrected read counts of the individual samples using Seurat version 4.3.0 (Hao et al., 2021) in R (version 4.2.1) (R: A language and environment for statistical computing. R Foundation for Statistical Computing). For data quality control, we followed the distributor’s outline. Cells were excluded when number of unique genes was <100 (nFeature_RNA) and/or when total number or RNA molecules was <1000 (nCount_RNA) and/or when percentage of mitochondrial genes was >25% (percent.mt).

Normalization and correction for cell cycle, mitochondrial and ribosomal gene percentage was performed using SCTransform. For both LCs and T cells separately, individual plates were integrated using the cca-based algorithm to find integration anchors in Seurat or using Harmony (Korsunsky et al., 2019), respectively. We calculated the Principal Components (RunPCA) and clustered the cells using Uniform Manifold Approximation (UMAP) (RunUMAP; FindNeighbors; 30 dimensions; FindClusters resolution of 0.4 for LCs and 0.8 for T cells). Log-normalized and scaled RNA counts were used for visualization.

### Statistical analysis of Single Cell RNA sequencing data

Testing for differential gene expression was performed using DESeq2 (Love et al., 2014) in the FindMarkers function in Seurat, where *P*-adjusted value < 0.05 was considered statistically significant. Gene signature scores were calculated using Seurat’s AddModuleScore function, based on the RNA assay. The compositions of these gene sets are listed in Table S9. Pathway analysis for the LC clusters was performed using the Gene Set Enrichment Gene Ontology (gseGO) function in clusterProfiler in R (Wu et al., 2021), which deploys all pathways available in the Gene Ontology knowledgebase (Ashburner et al., 2000, Gene Ontology et al., 2023). The Gene Set Enrichment Analysis (GSEA) function in R (Subramanian A., 2005), which deployed all pathways available in the Pathway Interaction Database (Schaefer et al., 2009), was used for analyzing DEGs of AD LCs versus healthy control LCs. A permutation test (scProportionTest, n permutations = 10,000) was used to determine statistical difference in cluster distributions between AD patients and healthy controls (Miller et al., 2021).

## Data availability

The RNA sequencing data that was obtained during this study have been deposited in NCBI’s Gene Expression Omnibus (GEO) and are accessible through GEO series accession number GSE266239.

## Acknowledgements

This work was supported by an Infection & Immunity Boost Grant from UMC Utrecht to F.v.W., M.d.B. and N.v.S. and by the DRESSCODE project (project number 09150181910001) of the Vici Talent program to N.v.S., which is financed by ZonMW.

## Author contributions

Conceptualization: F.v.W., N.v.S. Data curation: S.T., M.v.d.W., L.v.d.G. Formal Analysis: S.T., M.v.d.W. Funding acquisition: F.v.W., N.v.S., M.d.B. Investigation: S.T., M.v.d.W. Methodology: S.T., M.v.d.W., C.B. Resources: F.v.W., N.v.S., M.d.B., L.v.d.G. Software: S.T., M.v.d.W., E.S. Supervision: F.v.W., N.v.S., Y.P. Writing – original draft: S.T. Writing – review and editing: F.v.W., N.v.S., Y.P., A.H. All authors read and contributed to the final manuscript.

## Abbreviations used

AD: Atopic Dermatitis
*S. aureus*: Staphylococcus aureus
LCs: Langerhans cells
Th2 cell: T helper 2 cell
β-GlcNAc: β-linked-N-acetylglucosamine
WTA: wall teichoic acid
EASI: Eczema Area and Severity Index
FcεR1: high-affinity IgE receptor
HSP: heat shock protein
DEG: Differentially Expressed Gene
GSEA: Gene Set Enrichment Analysis.

