## Supplementary information for "Single Cell Sequencing of Human Langerhans Cells Identifies Altered Gene Expression Profiles in Patients with Atopic Dermatitis"

### Supporting Data

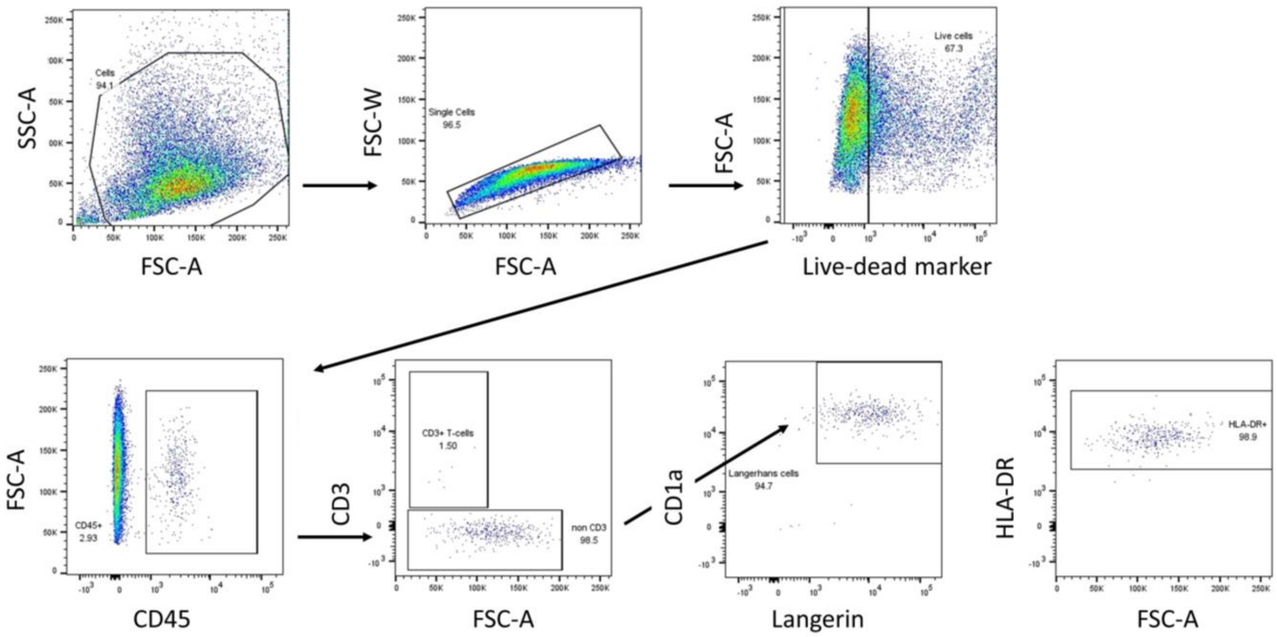


**Figure S1 Gating strategy for sorting LCs from skin epidermis**
Scatter plots showing the gating strategy used for fluorescence-activated cell sorting (FACS), for single Langerhans cells from human epidermis as live, CD45$^+$CD207$^+$CD1a$^+$cells. Plots were extracted from Flow Jo (version 10.9.0).

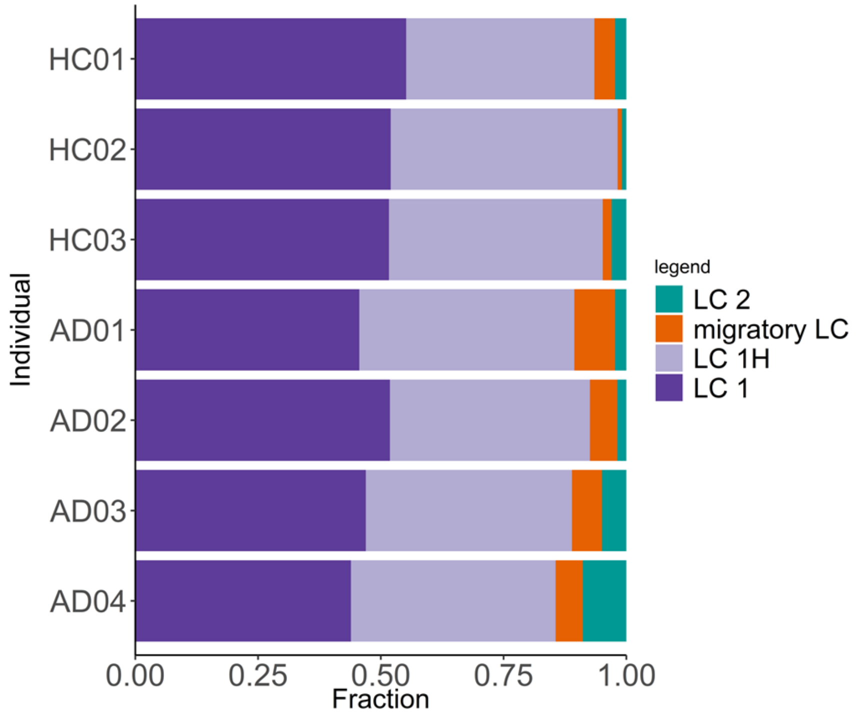


**Figure S2 Abundance LC clusters per individual**
Proportions of LC subsets present in each individual AD patient and HC, detected through single-cell RNA sequencing.

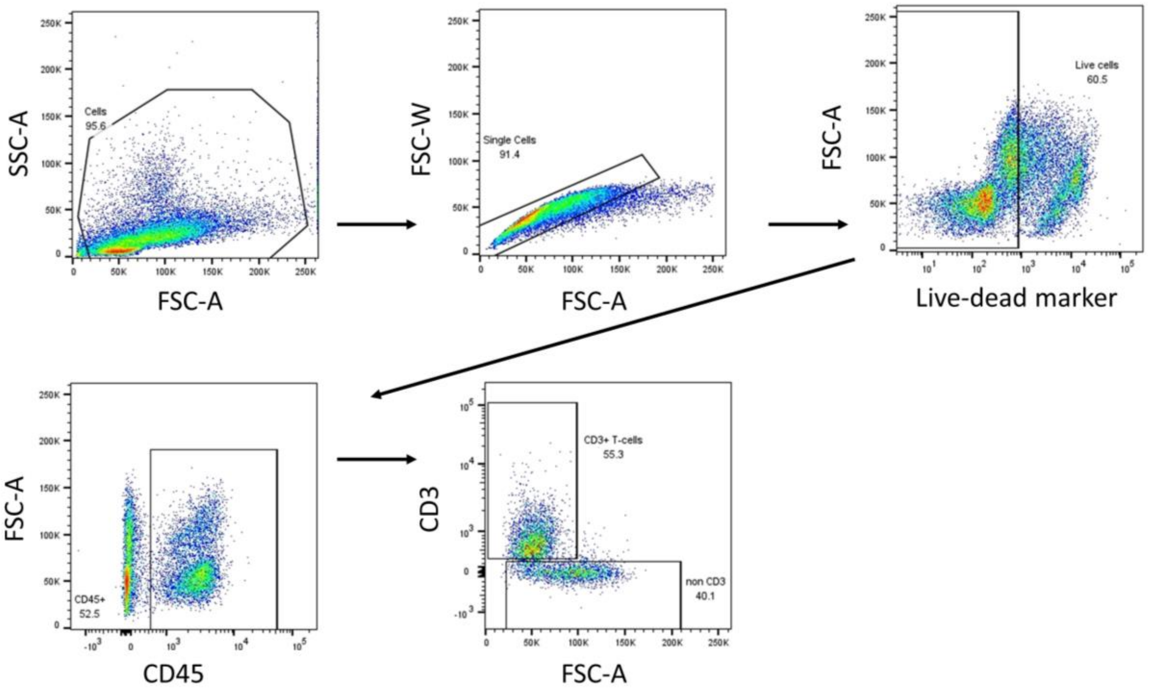


**Figure S3 Gating strategy for sorting CD3+ T cells from the dermis**

Scatter plots showing the gating strategy used for fluorescence-activated cell sorting (FACS), for single T cells from human dermis as live, $CD45^{+}CD3^{+}$ cells. Plots were extracted from Flow Jo (version 10.9.0).

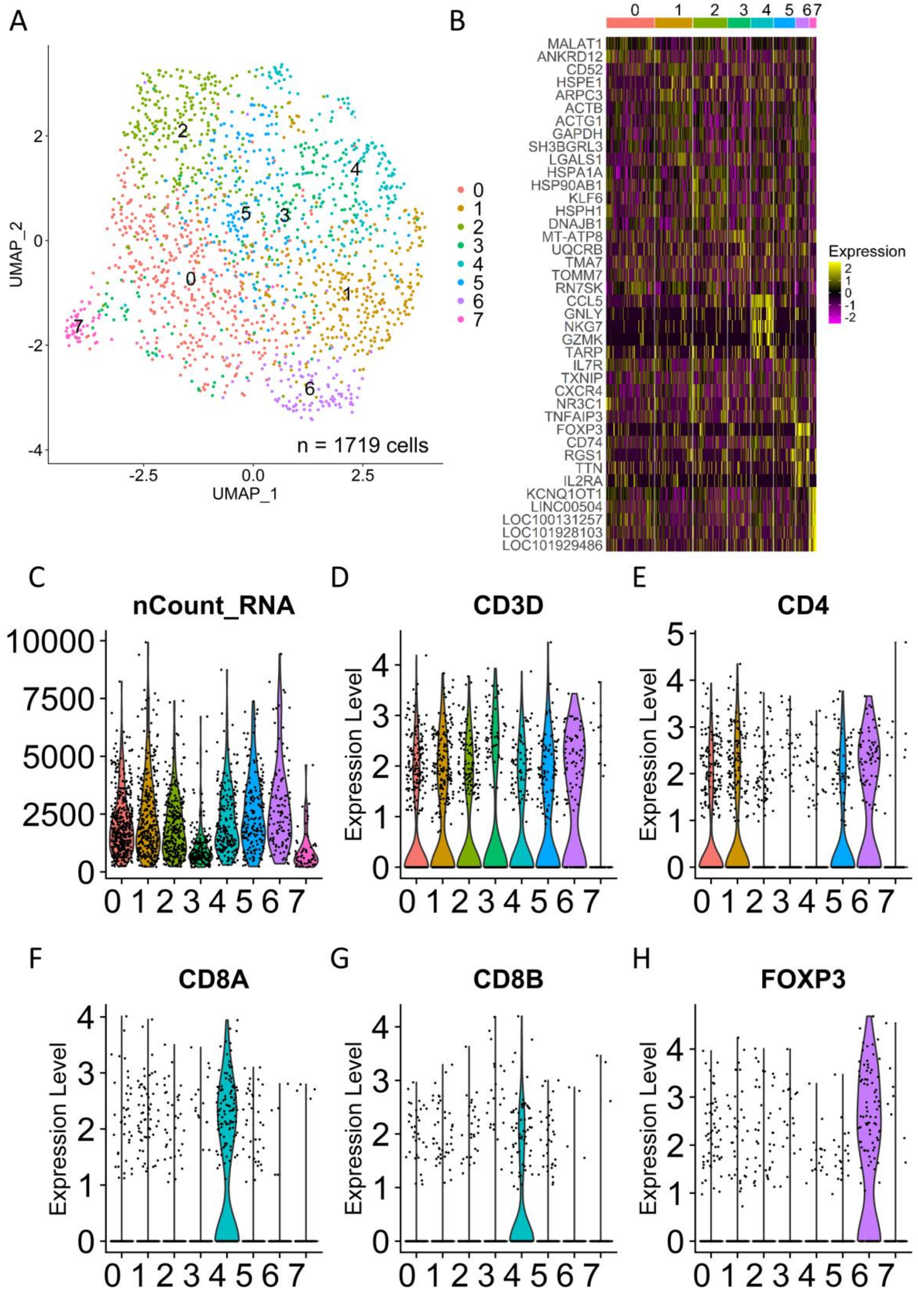

A
B
C
D
E
F
G
H
UMAP_1
UMAP_2
n = 1719 cells
Expression
MALAT1
ANKRD12
CD52
HSPE1
ARPC3
ACTB
ACTG1
GAPDH
SH3BGRL3
LGALS1
HSPA1A
HSP90AB1
KLF6
HSPH1
DNAJB1
MT-ATP8
UQCRB
TMA7
TOMM7
RN7SK
CCL5
GNLY
NKG7
GZMK
TARP
IL7R
TXNIP
CXCR4
NR3C1
TNFAIP3
FOXP3
CD74
RGS1
TTN
IL2RA
KCNQ1OT1
LINC00504
LOC100131257
LOC101928103
LOC101929486
nCount_RNA
CD3D
CD4
CD8A
CD8B
FOXP3
Expression Level

**Figure S4 Initial T cell clustering**

(A) UMAP plot of 1,719 individual CD3+ T cells from healthy and AD dermis, colored by cluster. (B) Heatmap of top 5 upregulated DEGs per cluster. (C) Volcano plot indicating total RNA molecules (nCount_RNA) per cell stratified per cluster. (D) Expression of CD3D gene per cell indicated by cluster (E)-(H) same as in (D) but for CD4 , CD8A. CD8B and FOXP3, respectively.

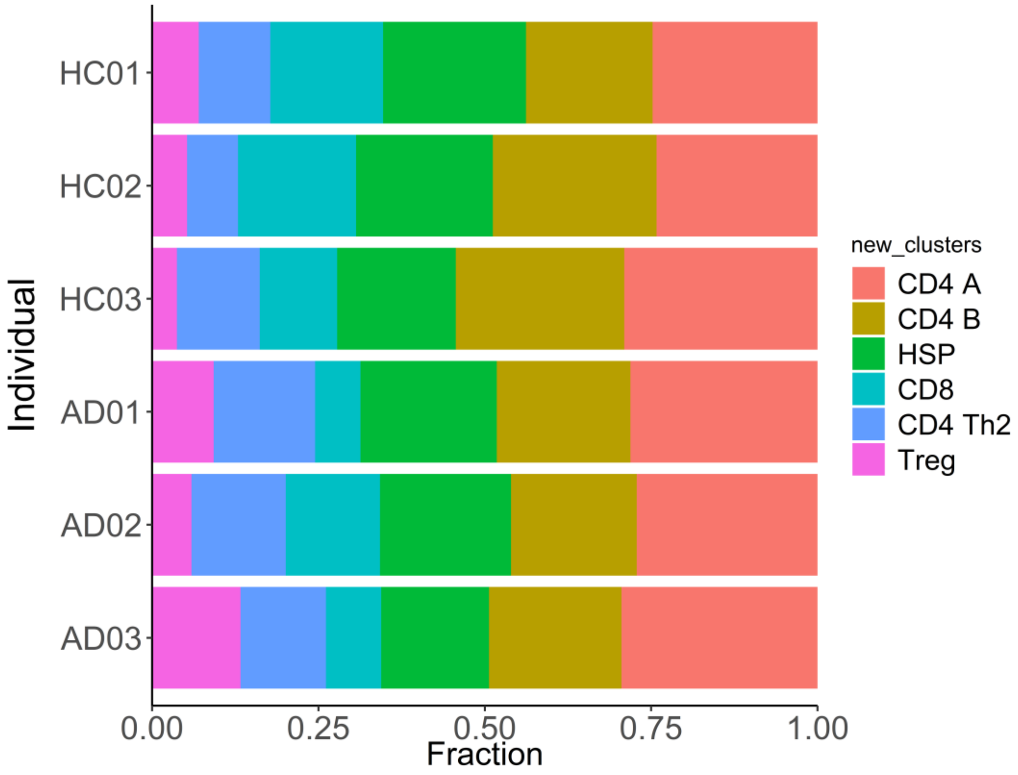


**Figure S5 Abundance T cell clusters per individual**
Proportions of T cell subsets present in each individual AD patient and HC, detected through single-cell RNA sequencing.

**Table S1. Isolated cell numbers per individual after quality control**

| Individual | Langerhans cells | T cells |
| --- | --- | --- |
| AD01 | 217 | 249 |
| AD02 | 108 | 254 |
| AD03 | 262 | 241 |
| AD04 | 271 | NA |
| HC02 | 261 | 242 |
| HC03 | 225 | 248 |
| HC04 | 333 | 241 |
| | **1,677** | **1,475** |

**Table S5 Curated genesets used in LC analysis**

| Geneset | Genes | References |
|---|---|---|
| FcεR signaling | FCER1A, FCER1G, ALOX5AP, ALOX5, BTK, JUN, FOS, CBL, CBLB, HCLS1, LCP2, NFATC2, PDHB, PLCB2, PLCB3, PLCG1, PLCG2, PLD2, PTPN6, PTPN11 | BioCarta; FC epsilon receptor signaling<br>FCER1A harmonizome 3.0<br>Kegg HSA04664 |
| Prostaglandin and leukotrienes biosynthesis | PLA2G4A, PTGS2, AKR1C3, PTGIS, TBXAS1, PTGDR2, PTGER3, PTGIR, ALOX5, ALOX5AP, ALOX15, NOP9, CYSLTR1, ABCC4, ANXA2, S100A10, S100A6 | Honda and kabashima 2019<br>WP98 wikipathways |
| C-type lectin receptors and signaling | CLEC10A, CLEC7A, CD207, MRC1, CD209, LY75, CLEC1A, CLEC4A, PYCARD, JUN, SYK, CALM1, PTGS2 | Figdor 2002<br>Liu 2021<br>Kegg c-type lectin signaling HSA04625 |

**Table S8 Primers used for detection of *tar*-glycosyltransferases**

| ***tar* gene** | **Forward primer** | **Reverse primer** |
|---|---|---|
| *tarS* | GTGAACATATGAGTAGTGCGTA | CATAATGTCCTTCGCCAATCAT |
| *tarM* | GGGATACCCATATATTTCAAGG | CAATTCGCTTCGTTGGTACCATTG |
| *tarP* | GTAAGTGTTATAATGCCAACATT | TATCAGCTTTCGCTACATTTCC |

**Table S9 Fluorescent dyes and antibodies used for cell sorting**

| Target | Clone | Fluorochrome | Manufacturer |
|---|---|---|---|
| Viability | Fixable Viability Dye | eFluor 506 | Invitrogen (65-2860-40) |
| CD45 | HI130 | BV711 | BioLegend |
| CD1a | HI149 | APC | Becton Dickinson |
| HLA-DR | G46-6 | BV605 | Becton Dickinson |
| CD207 | DCGM4 | PE | Beckman Coulter |
| CD8 | SK1 | PE-Cy7 | Sony Biotechnology |
| CD3 | UCHT1 | AF700 | BioLegend |
| CD4 | RPA-T4 | APC-Cy7 | BioLegend |
